# Protection from Killed Whole-Cell Cholera Vaccines: A Systematic Review and Meta-Analysis

**DOI:** 10.1101/2024.08.13.24311930

**Authors:** Hanmeng Xu, Amanda Tiffany, Francisco Luquero, Suman Kanungo, Godfrey Bwire, Firdausi Qadri, Daniela Garone, Louise C Ivers, Elizabeth C Lee, Espoir Bwenge Malembaka, Vincent Mendibourne, Malika Bouhenia, Lucy Breakwell, Andrew S Azman the Global Taskforce for Cholera Control Oral Vaccination Working Group

## Abstract

**Background:** Killed whole-cell oral cholera vaccines (kOCVs) are a standard prevention and control measure in cholera endemic areas, outbreaks, and humanitarian emergencies. Recently, new evidence has emerged and the ways in which the vaccines are used have changed. An updated synthesis of evidence on kOCV protection, is needed.

**Methods:** We systematically searched for randomized trials and observational studies that reported estimates of protection against confirmed cholera conferred by kOCVs. Eligible studies in English, French, Spanish or Chinese published through March 8, 2024, were included. Data on efficacy and effectiveness were extracted as were the number of doses, duration of follow-up, and age group. Efficacy and effectiveness estimates were summarized separately using random effects models to estimate protection by time since vaccination; meta-regression models were used to estimate protection, by dose, as a function of time since vaccination.

**Findings:** Twenty-three publications from five randomized controlled trials and ten observational studies were included. Average two-dose efficacy one-year post-vaccination was 55% (95%CI: 46-62%), declining to 44% (95%CI: 25-59%) four years post-vaccination. Average two-dose effectiveness was 69% (95%CI: 58-78%) one-year post-vaccination declining to 47% (95%CI: 9-70%) four years post-vaccination. Only one randomized trial assessed one-dose efficacy and found sustained protection for two years (52%; 95%CI 8-75%). Average one-dose effectiveness one year after vaccination was 60% (95%CI: 51-68%) and 47% (95%CI: 34-58%) after two years. Through age-group specific meta-analysis average 2-dose efficacy in children under five years old was half that of older individuals.

**Interpretation:** Two doses of kOCV provide protection against medically-attended cholera for at least four years post-vaccination. One-dose of kOCV provides protection for at least two years post-vaccination, but wanes faster than that of two doses. Children under five are less protected by kOCVs compared to those 5 years and older regardless of the number of doses received.

**Funding:** Bill and Melinda Gates Foundation

## Introduction

Killed whole-cell oral cholera vaccines (kOCV) are part of the standard cholera control and prevention package often used in combination with fundamental interventions like water, sanitation and hygiene improvements. They have been used reactively to control outbreaks where the short-term vaccine protection is most critical, and preemptively in areas with endemic cholera where longer term vaccine protection is key.

The current generation of kOCVs underwent the first clinical trials in the 1980’s with two versions tested, a simple kOCV and the other a kOCV with a recombinant B-subunit of the cholera toxin.^1^ Both went on to be developed, but today, vaccines with the B-subunit are used only for those traveling to cholera endemic areas. All vaccines used in public health programs in cholera-affected countries, including those used preventively and for outbreak response through the global stockpile, are WHO approved (prequalified) kOCVs, including both primary serotypes of 7th pandemic *Vibrio cholerae* O1. Four biologically similar kOCVs have been approved by the WHO for use through the stockpile: Shanchol in 2011 (Sanofi Pasteur), Euvichol in 2015 (EuBiologics), Euvichol-Plus in 2017 (EuBiologics), and Euvichol-S in 2024 (EuBiologics).

Clinical trials and observational studies in several cholera endemic countries like Bangladesh, Haiti, and India have shown that kOCVs are safe and immunogenic, though estimates of the level of protection vary widely across studies.^2–5^ Vaccine-derived protection wanes over time, though precise estimates of the rate of waning across different epidemiologic settings are lacking. A systematic review and meta-analysis of kOCV protection against cholera published in 2017 found a pooled estimate of two-dose efficacy of 58% and field effectiveness of 76%, with protection from two doses lasting at least three years.^6^ However, few studies included follow-up beyond two years after vaccination. Protection in children under 5 was lower, though analyses did not control for differences in epidemiologic setting and few studies provided age-stratified estimates.

Countries can access kOCV for outbreak response and preventive campaigns, with most cholera-affected countries eligible to have vaccine and campaign operational costs covered by Gavi, the Vaccine Alliance. Since 2021, demand for kOCVs has far outstripped global supply.^7,8^ Consequently, the International Coordinating Group, which oversees allocation of the global stockpile for outbreak response, temporarily suspended the standard two-dose regimen of kOCVs for outbreak response and replaced it with a single dose strategy in October 2022, which continues at the time of writing.^9,10^ In this context, no two-dose preventive campaigns were conducted in 2023. Furthermore, in 2023, Sanofi ceased production of Shanchol, the vaccine for which most evidence on protection has been generated. At the time of writing, three kOCVs are available through the stockpile; Euvichol/Euvichol-Plus, which have the same strains of killed bacteria as Shanchol, and Euvichol-S, which has only a subset of the other vaccines’ strains.^11,12^ All three of these vaccines were licensed on the basis of immunological studies, with no clinical end-points.

Due to new data on protection from the WHO-approved kOCVs, changes in kOCV use and increasing global kOCV demand, there is a need to review the available data on protection by dose, population and epidemiologic setting to inform outbreak response and revaccination policies. In this study, we update a previous systematic review and meta-analysis to provide a more detailed view on the protection provided by WHO-approved kOCVs as a function of the number of doses received, duration of follow-up, and age.

## Methods

### Literature search and data abstraction

We searched PubMed, Embase, Scopus, ISI Web of Science, and the Cochrane Review Library database for literature published through March 8, 2024, using the search terms used in the previously published systematic review,^6^ but restricted the search to literature published from January 1, 2016, the end date of the previous systematic review search (Table S1). Records identified by the search terms were imported to a web-based screening tool (https://www.covidence.org/) where duplicates were automatically detected and removed. We also consulted several experts through the Global Task Force on Cholera Control to identify potential publications missed by our search strategy.

Titles and abstracts were each independently screened by two of three reviewers (HX, ASA, AT) using the same inclusion and exclusion criteria used in the previous review;^6^ conflicts were resolved by a third reviewer or by discussion. We included clinical trials or observational studies published in English, Spanish, French or Chinese that enrolled medically attended confirmed cholera cases (based on at least one diagnostic test for *V. cholerae* O1/O139) to understand the efficacy or effectiveness of kOCV. Both new studies and continued follow-up of previous studies were included. Publications that had been included in the previous review were marked as duplicates and excluded from the new search. We compiled the results identified during the new search and those from the previous review for the analysis.

Two reviewers (HX and AT) independently assessed the risk of bias of each included study (both new and old) using the Newcastle-Ottawa Scale for the observational studies and the Cochrane Collaboration’s tool for randomized clinical trials (RCTs). Conflicts were resolved through discussion or by a third reviewer. The assessment was based only on the methods described in the publications. If one study had more than one publication for different follow-up periods, the information presented in the methods sections of all publications were considered while assessing the risk of bias.

During full text review, we extracted data on the study setting, target population, study type, vaccine, dosing regimen, case confirmation method, method(s) of vaccination status ascertainment, estimates of vaccine protection (vaccine efficacy from randomized trials and effectiveness from observational studies), and measures of uncertainty. When available, we extracted multiple estimates of vaccine protection including those disaggregated by age, time since vaccination and number of doses received. Data from studies reporting estimates of efficacy or effectiveness of kOCVs with recombinant B-subunit were extracted but not included in the analyses as their use is currently restricted to travelers. Only those estimates that were reported in manuscripts were extracted, and we did not attempt to use estimate presented in the manuscript to calculate new estimates of protection.

### Data analysis

Similar to the previous review by Bi et al., we used the reported point estimates of vaccine efficacy or effectiveness and 95% confidence intervals (CIs) to calculate the standard error of each. For estimates with only one-sided CIs, two-sided 95% CIs were reconstructed.^6,13^ All estimates were extracted as reported by the authors. As kOCV protection wanes, our analyses focused on estimating effectiveness and efficacy as a function of the time since vaccination, first through time-bin stratified analyses and then using meta-regression.

In stratified analyses, we grouped estimates of vaccine protection based on the mid-point of the follow-up period (e.g., 0-12, 12-24, 24-36, 36-48, and 48-60 months post-vaccination). For each time bin, we estimated a pooled mean vaccine efficacy/effectiveness using a random-effects model with an empirical Bayes estimator for the between-study variance and assessed heterogeneity using the I^2^ statistic.^14,15^

The stratified estimates of protection synthesize estimates from slightly different follow-up periods (i.e., within the 0-12 month bin there may be estimates pertaining only to the first three months post vaccination and others representing the entire period) and estimates are not constrained to only decrease or remain constant over time. Thus, for the primary analyses we used a meta-regression approach to estimate protection as a function of time since vaccination, incorporating all estimates of protection with their exact time range post-vaccination (e.g., “15 to 22 months post vaccination”). We fit mixed-effects meta-regression models with the natural logarithm of time since vaccination as the primary fixed effect and between study variance estimated with a restricted maximum likelihood estimator as implemented in the metafor package in R.^15^ We estimate and present 95% confidence intervals for the mean protection over time in addition to the 95% prediction intervals, which illustrate the expected variability in future studies across different settings. We explored alternative meta-regression models with different transformations of time since infection and models with random slopes and compared support for these using Akaike Information Criterion (AICc). We used the midpoint of the period pertaining to each estimate in the model and fit separate models by dose and study type (effectiveness and efficacy). We explored the influence of each data point on the estimated waning curves through conducting leave-one-out analyses.

To explore vaccine protection in those under 5 years of age compared to 5 years of age and older, age-stratified efficacy/effectiveness estimates were extracted. Estimates by age group were pooled and the average vaccine efficacy/effectiveness were estimated by number of doses and age group. If a study reported both estimates for under 5 years and 5 years and older, we estimated the relative vaccine effect ratio, calculated as vaccine effect in those under 5 years divided by vaccine effect in those 5 years and older.

All the analyses were performed in R version 4.3.^16^ The original 2016 review was pre-registered in the PROSPERO database under CRD42016048232.

Role of the funding source

The funder of the study had no role in the study design, data collection, data analysis, data interpretation, or writing of the report.

## Results

We identified 8,205 records published online through March 8, 2024 in our searches, including 6,224 identified in the previous review and 1,981 identified in our new search (after January 1, 2016). Of these, 53 records were eligible for full text review. One additional full-text review-eligible publication was identified from a reference review of identified publications.^17^ Twenty-three publications met the inclusion criteria for data abstraction, which included estimates from five randomized controlled trials (13 publications^1–4,18–26^) and ten observational studies (10 publications^5,17,27–34^) (Figure 1, S1, Table 1, 2).

**Figure 1.**
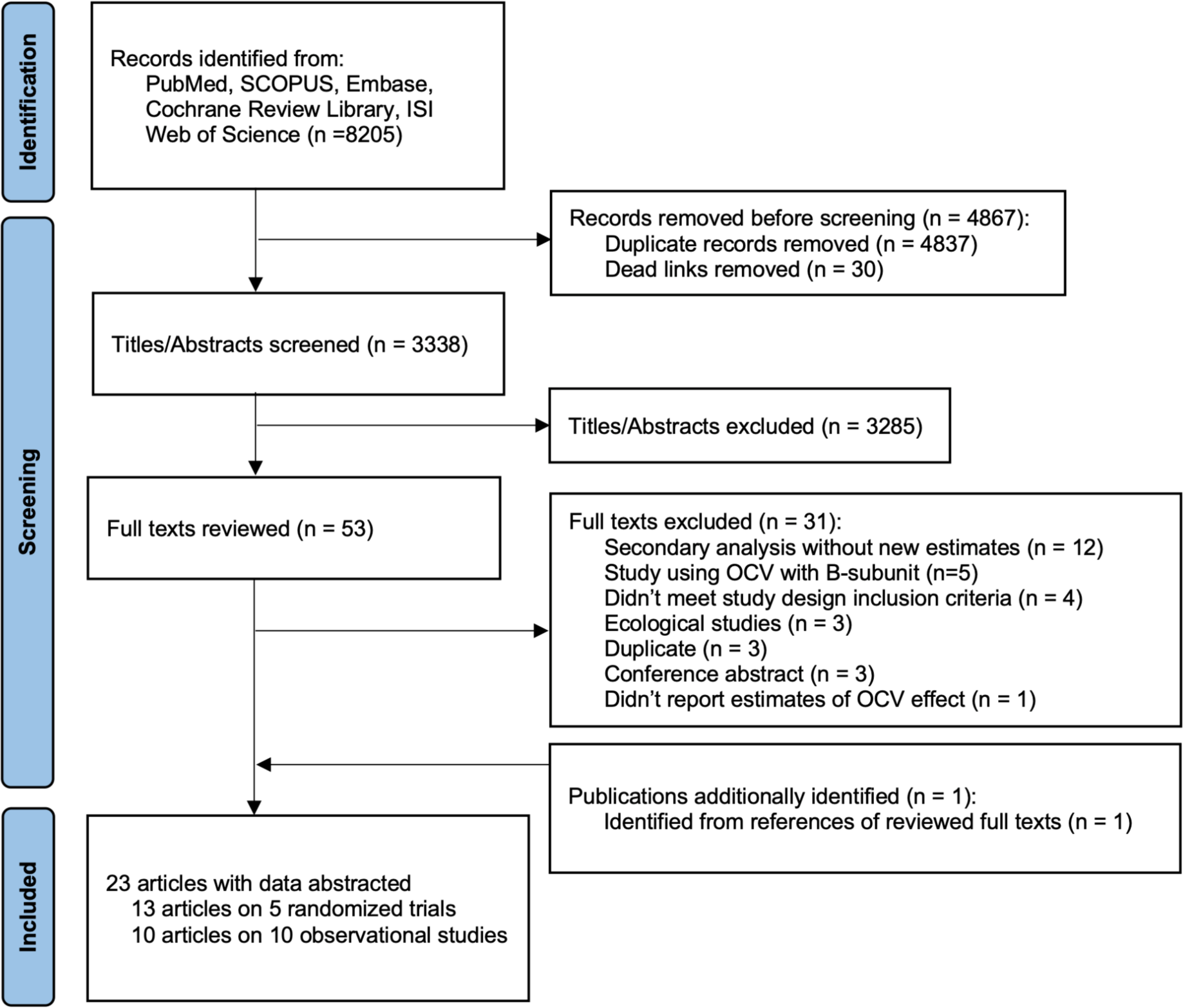
PRISMA flow chart of the record screening process. This flowchart illustrates the screening process for the combined dataset of records identified from the 2016 and 2023/2024 searches.

**Table 1.** Overview of clinical trials leading to estimates of vaccine efficacy that met the inclusion criteria.

| Study initiation year** | Study location | Study design | Vaccine | Post-vaccination follow-up duration | Number of doses for primary outcome | References |
| --- | --- | --- | --- | --- | --- | --- |
| 1985 | Matlab, Bangladesh | Individually randomized controlled trial | WC*** | 48 months | Three | 1,2,20–22 |
| 1992 | Hue, Vietnam | Household randomized trial | WC*** | 10 months | Two | 24 |
| 2006 | Kolkata, India | Cluster-randomized controlled trial | Shanchol | 60 months | Two | 4,18,19 |
| 2011 | Dhaka, Bangladesh | Cluster-randomized trial | Shanchol | 48 months* | Two | 3,26 |
| 2014 | Dhaka, Bangladesh | Cluster-randomized trial | Shanchol | 24 months* | One | 23,25 |

\*New follow-up estimates not included in Bi et al.
\*\* Year when the first vaccination occurred
\*\*\* precursor to Dukoral (without B subunit) and Shanchol with similar antigenic composition.

**Table 2.**
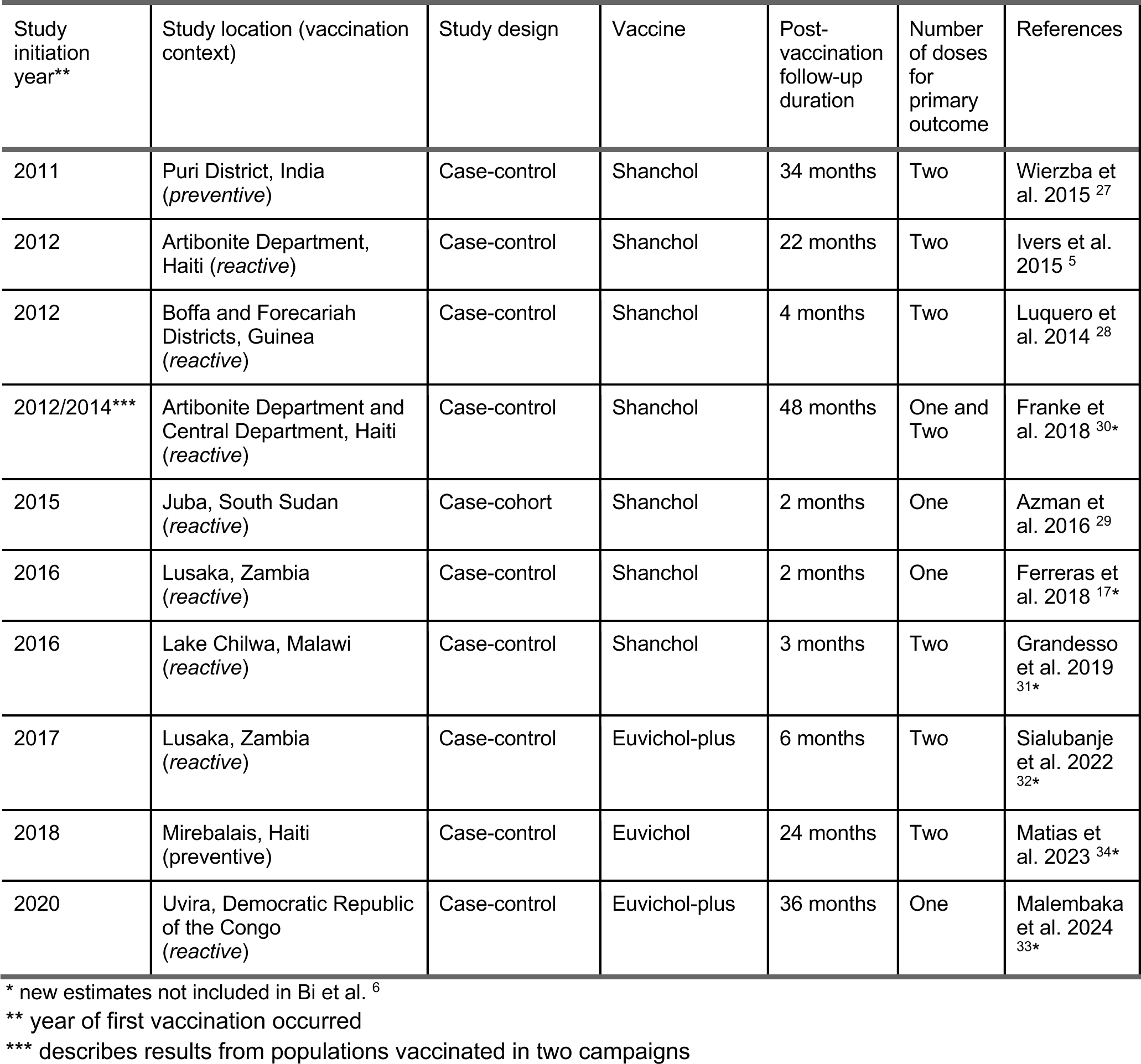
Overview of effectiveness studies that met the inclusion criteria.

The five randomized controlled trials, yielding estimates of vaccine efficacy, included were conducted in Kolkata (India), Matlab (Bangladesh), Dhaka (Bangladesh), and Hue (Vietnam), the earliest trial starting in 1985 in Matlab, and the most recent starting in 2014 in Dhaka. One trial reported efficacy estimates for a three-dose regimen, three for a two-dose regimen, and one for a one-dose regimen. The post-vaccination follow-up time ranged from 10 months to 5 years after vaccination for two- or three-dose trials, and 2 years for the single one-dose trial. The two early trials, in Matlab and Hue, used vaccine formulations that were precursors to the current kOCVs. The three trials in Kolkata and Dhaka used Shanchol. None of these trials used Euvichol or Euvichol-Plus. Four trials reported that case finding was done only through passive surveillance in clinics where cholera cases were identified and treated, while the earliest trial in Matlab also conducted active community-based case finding.^1^ Outcomes in all trials were based on culture confirmation. Two studies had a low risk of bias across all study quality domains,^19,23^ one had low risk across all but two study domains where the risk was “unclear”.^1^ The remaining two studies had a high risk of bias in a few domains, including those related to blinding of participants, outcomes and allocation concealment (Figure S2).^3,24^

The ten observational studies included (nine case-control studies and one case-cohort study) had a wider geographic range than the efficacy studies with six from sub-Saharan Africa, one from Asia, and three from the Caribbean. Seven were initiated after vaccination campaigns conducted in response to an outbreak and three were conducted preemptively in endemic areas including Puri (India), Artibonite and Central Department (Haiti), and Uvira (DRC). Two studies were conducted after the use of a one-dose emergency vaccination campaign.^17,29^ The post-vaccination follow-up period ranged from 3 months to 4 years for two-dose estimates, and 2 months to 4 years for one-dose estimates. Seven studies used Shanchol, one study used Euvichol and two studies used Euvichol-Plus. All observational studies identified cases through passive clinical surveillance.

Four studies used culture alone to classify cholera cases in their main analyses,^5,27,30,32^ two used PCR alone,^31,34^ one used culture and PCR,^17^ and three used a combination of PCR, culture and rapid diagnostic tests.^28,29,33^ Nine out of ten observational studies ascertained vaccination status based on self-reporting and reference to vaccination card if available, and only one study relied solely on electronic vaccination registry.^27^ Availability of vaccination card among cases who reported to have been vaccinated varied across studies, ranging from 0% to 82%, with a median of 50%. All ten observational studies had a low risk of selection bias, one of these had a low risk of bias related to comparability. All nine case-control studies did not report adequate data on non-response rates (Table S2).

### Two-dose protection

We identified sixteen estimates of efficacy (from five trials) and thirteen estimates of effectiveness (from eight observational studies) for two-doses of kOCV (Figure 2A, 2B, Table S3). Five efficacy estimates pertained to protection 0-12 months post-vaccination, four to protection 12-24 months post-vaccination, three to 24-36 months post-vaccination, three to 36-48 months post-vaccination and one to 48-60 months post-vaccination. Similarly, five effectiveness estimates pertained to protection 0-12 months post-vaccination, four to 12-24 months post-vaccination, three to 24-36 months post-vaccination and one to 36-48 months post-vaccination (Table S3). Within the first year (0-12 months) after vaccination, estimates of efficacy ranged from 40% (95% CI: -10–67%) to 66% (95% CI: 46-79%), with effectiveness estimates ranging from 81% (95%CI: 72–84%) to 87% (95% CI: 32-98%). In the second year (12-24 months) after vaccination, efficacy estimates ranged from 57% (95% CI: 42–69%) to 72% (95% CI: 42-87%), and effectiveness estimates ranged from 58% (95% CI: 27–76%) to 69% (95% CI: -71-94%). In the third year (24-36 months) after vaccination, efficacy estimates ranged from 25% (95% CI: -13–51%) to 57% (95%CI: 26–75%) and effectiveness estimates ranged from 25% (95% CI: -19–52%) to 73% (95% CI: 30–90%). In the fourth year (36–48 months) after vaccination, efficacy estimates ranged from less than zero to 60% (95% CI: 33– 76%) and the one study estimated 94% (95% CI: 56–99) effectiveness (Figure 2A,2B).

**Figure 2.**
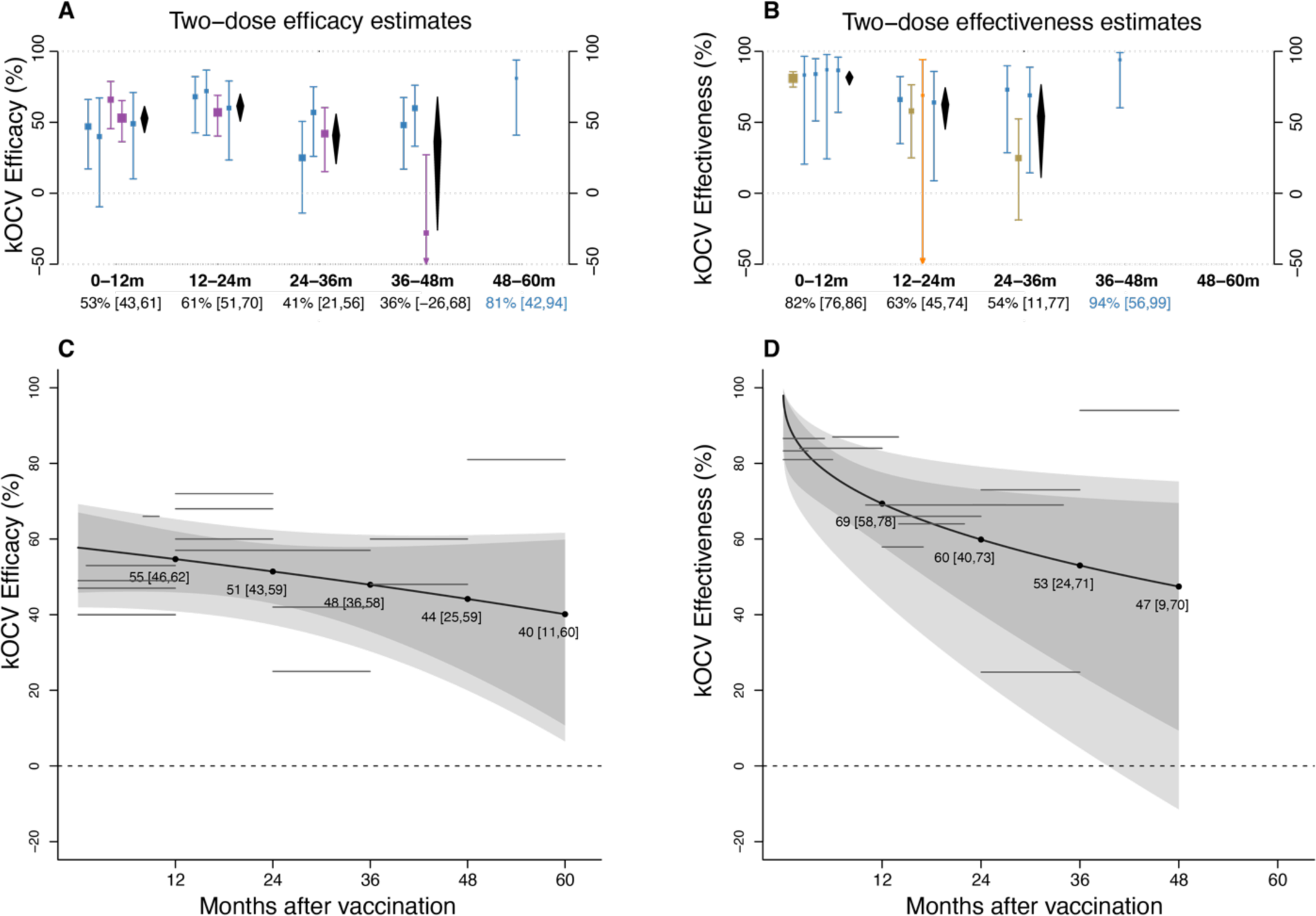
Stratified and meta-regression estimates of the efficacy and effectiveness of two doses of killed whole-cell OCV (kOCV) as a function of time (months) since vaccination. The upper panels illustrate stratified estimates of efficacy (A) and effectiveness (B) by time since vaccination bin (0-12, 12-24, 24-36, 36-48, 48-60 months after vaccination). The trial conducted in Matlab, Bangladesh was using three doses of OCV but was also included in this figure 20. Estimates are grouped into the five follow-up duration categories by the midpoint of the time window during which the estimate was measured. Bars and squares show 95% confidence intervals (CI) and point estimates of efficacy or effectiveness for each literature, colored by vaccine type (blue: “Shanchol; gold: Euvichol-plus; orange: Euvichol; purple: whole-cell vaccines used in early trials that were precursors of Shanchol vaccine). Diamonds in black show the estimated average efficacy or effectiveness and 95% CI by follow-up period, with numerical values shown at the bottom of the x-axis in black. If there is only one estimate in the follow-up period, the estimate from the study is presented on the x-axis. The bottom panels illustrate meta-regression results for average two-dose (C) efficacy and (D) effectiveness as a function of time since vaccination, with the shaded envelope representing the 95% confidence intervals and 95% prediction intervals (lighter region). The horizontal gray lines represent the data from the literature that were used to fit the meta-regression models, the length of the line indicates the duration of follow-up (months since vaccination). The line’s position on the y-axis marks the magnitude of the point estimate (%). The dashed horizontal line at y=0 denotes no protective effect (0%) of kOCV.

Given that protection wanes after vaccination, we fit meta-regression models to estimate the extent of two-dose protection over time (Figure 2C and 2D). Two-dose efficacy is 55% (95% CI: 46-62%) at 12 months post-vaccination, 51% (95% CI: 43–59%) at 24 months, 48% (95% CI: 36-58%) at 36 months, 44% (95% CI: 25–59%) at 48 months and 40% (95% CI 11-60%) at 60 months (Figure 2C). Using the same approach, effectiveness estimates are generally higher than efficacy estimates in the first two years (Figures 2C and 2D). Two-dose effectiveness is 69% (95%CI: 58-78%) at 12 months post-vaccination, 60% (95%CI: 40-73%) at 24 months, 53% (95%CI: 24-71%) at 36 months and 47% (95%CI: 9-70%) at 48 months. Leave-one-study-out meta-regression analyses illustrated that the two longest efficacy ^18,20^ and two longest effectiveness studies ^30,33^ influenced estimates of protection, especially in the fourth and fifth years after vaccination (Figure S3).

### One-Dose Protection

We identified four efficacy estimates (from one trial) and ten effectiveness estimates (from eight observational studies) reported for one-dose kOCV (Figure 3, Table S4). The four efficacy estimates pertained to 0-6, 6-12, 12-18, and 18-24 months post-vaccination (Table S4). Four effectiveness estimates pertained to the first 6 months post-vaccination, one for 6-12 months post-vaccination, three 12-18 months post-vaccination, and two 24-30 months post vaccination. One-dose efficacy was 58% (95% CI: 24–76%), 37% (95% CI: -20–67%), 62% (95% CI: 34–78%) and 67% (95% CI: 30–84%) for 0-6, 6-12, 12-18, and 18-24 months post vaccination, respectively. One-dose effectiveness estimates for 0-6 and 6-12 months post-vaccination ranged from 43% (95% CI: -84–82%) to 92% (95% CI: 66–98%). For 12-18 months post-vaccination, the three effectiveness estimates ranged from 40% (95% CI: -31–73%) to 67% (95% CI: -62-93%). The two one-dose effectiveness estimates pertaining to 24 to 30 months post-vaccination were 46% (95% CI: 26–60%) and 32% (95% CI: -318–89%).

**Figure 3.**
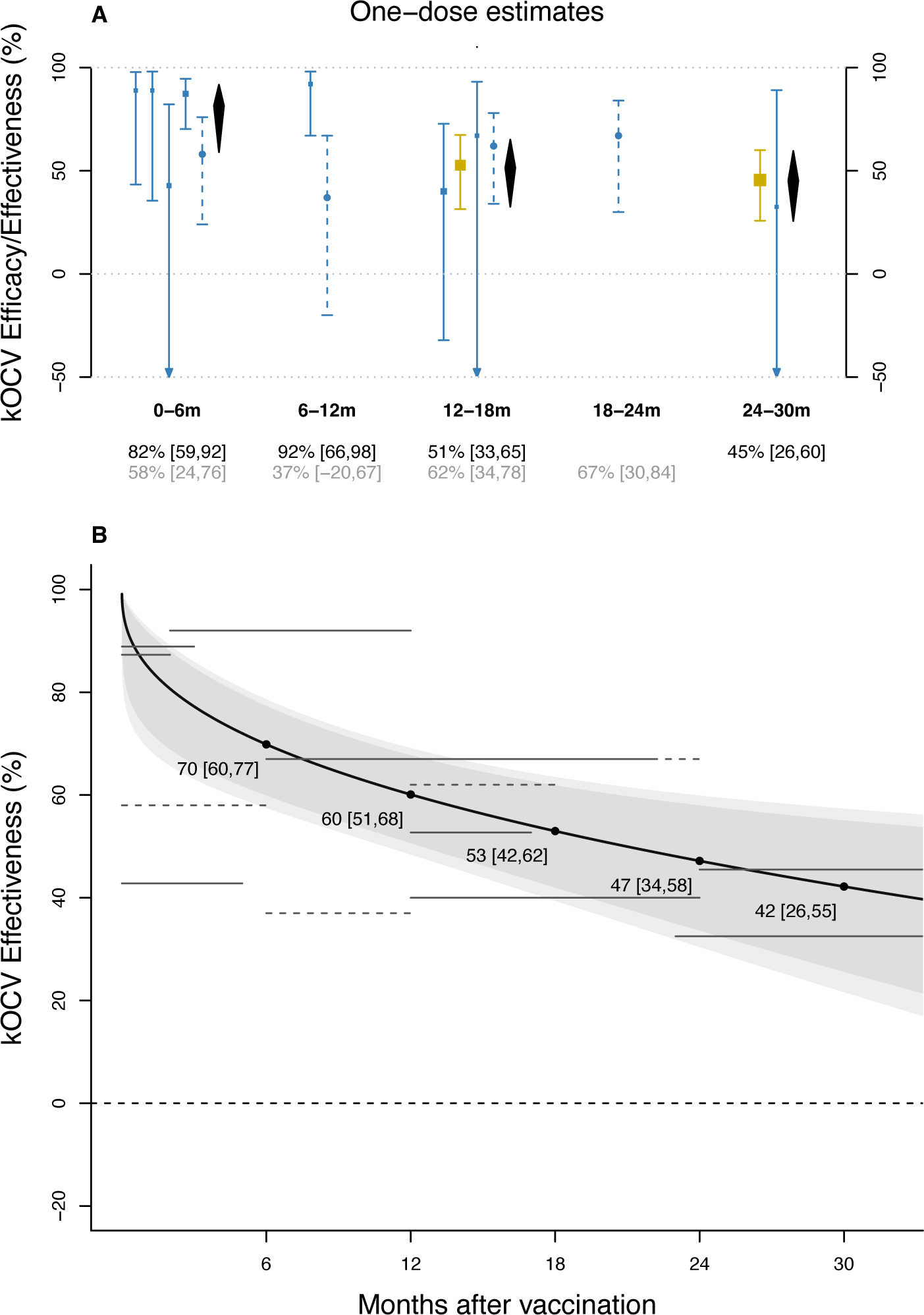
Stratified and meta-regression estimates of the efficacy and effectiveness of one dose of killed whole-cell OCV (kOCV) as a function of time since vaccination. The upper panel (A) illustrates stratified estimates of efficacy and effectiveness by 6-month time bins post-vaccination. Bars and squares show 95% confidence intervals (CI) and point estimates of efficacy (in dashed lines) or effectiveness (in solid lines), colored by vaccine type (blue: Shanchol; gold: Euvichol-Plus). Diamonds in black show the estimated average effectiveness and 95% CI by follow-up period, with numerical values shown at the bottom of the x-axis in black (first row). If there is only one effectiveness estimate in the follow-up period, the estimate from that study is presented. The second row of the x-axis label presents the efficacy estimates (in grey). The bottom panel (B) illustrates meta-regression results for average one-dose effectiveness as a function of time since vaccination, with the shaded envelope representing the 95% confidence intervals and 95% prediction intervals. The horizontal gray lines represent the data from the literature that were used to fit the meta-regression models, the length of the line indicates the duration of follow-up (months since vaccination), colored by study type (efficacy estimates: dashed; effectiveness studies: solid). The line’s position on the y-axis marks the magnitude of the point estimate (%). The dashed horizontal line at y = 0 denotes no protective effect (0%) of kOCV.

Estimates from meta-regression analyses for protection provided by one-dose of kOCV illustrate that effectiveness estimates within the first 12 months post vaccination are similar between one- and two-dose regimens, but one-dose protection appears to decay faster (Figures S4). We estimate that one-dose effectiveness 6 months post-vaccination is 70% (95% CI: 60–77%) and decreases to 60% (95% CI: 51–68%), 53% (95% CI: 42–62%), 47% (95% CI: 34–58%) and 42% (95%CI: 26–55) at 12, 18, 24 and 30 months, respectively. We estimated similar results for one-dose protection over time in leave-one-study-out analyses (Figure S5).

### Protection by age

Five trials and two observational studies that used two or three doses of kOCV reported age-stratified estimates (Table 3). Across all studies reporting age-stratified two-dose estimates, protection was consistently lower in children under 5 years old, except for one study in Vietnam that found similar point estimates between groups. Across studies with age-stratified estimates, the pooled two-dose efficacy for children under 5 years is 31% (95% CI: 14–45%, I^2^ = 0%), with a 36-month weighted mean follow-up period, compared with 62% (95% CI: 49–71%, I^2^ = 60%) for participants 5 years and older, across a 37-month weighted mean follow-up period (Figure S6).

**Table 3.**
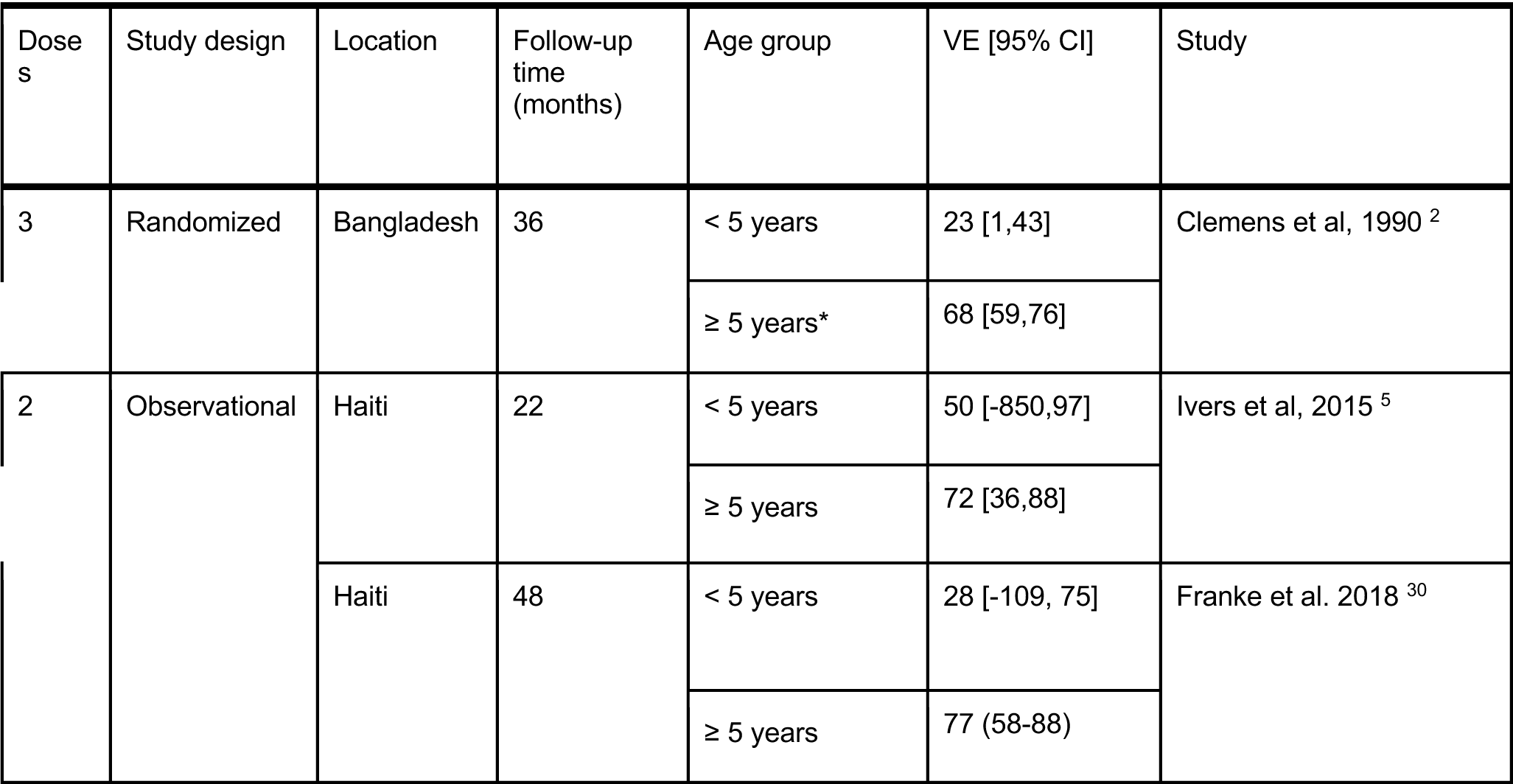
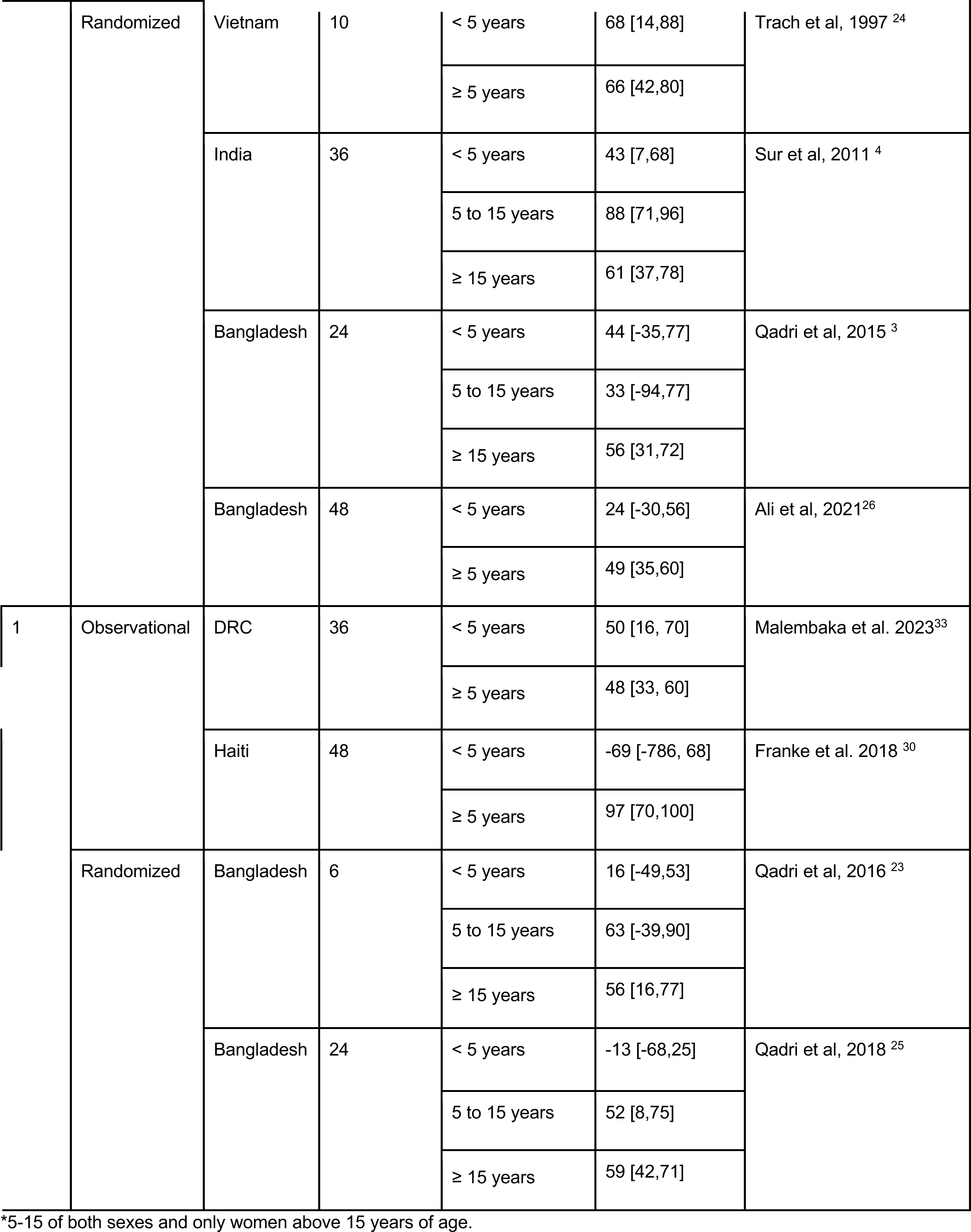
kOCV effectiveness and efficacy by age group.

Evidence for one-dose protection among young children continues to be limited. One randomized trial in Bangladesh and two case-control studies in the Democratic Republic of the Congo and Haiti reported estimates for children under 5 years old (Table 3, Figure S4). The randomized clinical trial in Bangladesh found that protection among children under 5 years at 6- and 24-months post-vaccination was 16% (95% CI: -49–53%) and -13% (95% CI: -68–25%). In comparison, protection among those 5 years and older was 57% (95% CI: 24–76%) and 58% (95% CI: 42–69%). In contrast, the case-control study in the Democratic Republic of the Congo found nearly identical estimates of protection across the two age groups, reporting 50% (95%CI: 16–70%) in children under 5 years and 48% (95%CI: 33–60%) in those 5 years and older during 12-36 months post-vaccination.

## Discussion

In this review, we summarize data from 5 randomized controlled trials and 10 observational studies covering 10 countries and find evidence that the substantial protection provided by the standard two-dose kOCV regimen lasts for at least four years after vaccination. Protection from a single-dose regimen was sustained at least over the first two years after vaccination though waning appeared to be faster than that of two doses (Figure S4). Evidence on one and two-dose protection is scant in populations with little to no historical exposure to *V. cholerae* O1. Despite the moderate-to-high estimates of vaccine protection, most studies have estimated substantially lower direct protection in children 1 to 4 years old when compared with the rest of the population.

Since the last systematic review and meta-analysis of kOCV protection was published in 2017, the use of cholera vaccines has increased substantially both in terms of the number of doses and the number of countries conducting campaigns. While 2022 and 2023 saw a shift to nearly exclusive use of the vaccine for outbreak response, cholera endemic countries are also planning for more sustained use of kOCV as part of a multi-year preventive program in the years to come.^35^ Several important questions on how to design vaccination programs remain: where to vaccinate, when to revaccinate, the number of doses to use (one versus two, especially if revaccinating) the optimal timing between doses, and whether recommendations should be age-group specific. The 2017 WHO position paper on cholera vaccines states that populations should not be re-vaccinated within a three-year period.^36^ Our results suggest that, at least in populations frequently exposed to *V. cholerae* O1, protection from two doses is sustained over this period with evidence from both randomized and observational studies demonstrating protection well beyond three years – in the 4th and 5th years after vaccination. Only short-term estimates of protection, within the first year after vaccination,^17,29^ are available from settings that had not reported cholera for several years, so the duration of vaccine protection in immunologically naive populations remains unclear. Additional data are needed on kOCV effectiveness in non-endemic settings with populations that have had little to no previous exposure to pandemic *V. cholerae*.

In 2022, the International Coordinating Group on Vaccine (ICG), the body that makes allocation decisions for the emergency stockpile of kOCVs, temporarily suspended use of two-dose regimens of kOCV due to the global kOCV shortage and the high demand.^10^ Our synthesis of evidence on one dose protection suggests that for outbreak control, where protection in the first year is key ^37^, this may be an appropriate dosing strategy. While evidence points towards one dose protection for at least two years in the general population, there are no data on one-dose protection in populations with little to no background exposure to pandemic *V. cholerae*. Results from Bangladesh suggest that one dose provides little to no protection to children under 5 years old ^23^, in contrast to a case-control study from DR Congo,^33^ that found similar levels of protection provided by kOCV across age groups in the first 2 years post-vaccination. The reasons for this discrepancy between studies are unclear, however, one potential explanation is that indirect effects from vaccinated household members in each study differed, with the individually-randomized trial less subject to the influence of household indirect effects.^38,39^

Our attempt to synthesize a diverse set of evidence on direct vaccine protection is not without limitations. In contrast to previous systematic reviews, we did not attempt to characterize protection by a single number, given that protection wanes over time (Figures 2 and 3). Instead, we provided time-stratified pooled estimates and used a meta-regression analysis to estimate expected protection as a function of time since vaccination. We did not have access to individual-level data from most published studies and had to rely on non-standardized, and sometimes coarse, time intervals for our meta-analyses. In our meta-regression we attributed each point estimate from the literature to the midpoint of the time interval since vaccination, though this should be assigned to a case-weighted summary of time since vaccination. While the vaccines used in the studies included in our main analyses were similar to one another, they do not have exactly the same composition. Most evidence comes from Shanchol, a vaccine that is no longer produced, with only 3 estimates from observational studies of Euvichol/Euvichol-Plus and no estimates related to Euvichol-S, the currently available kOCVs. We did not have enough data to reliably detect differences in protection between different vaccines, though the point estimates from Euvichol/Euvichol-Plus are generally consistent with prior evidence from other vaccines (Figure 2, Figure 3). As a sensitivity analysis, we refit meta-regression models excluding data from studies that used vaccines that came before Shanchol^20,24^, that are known to have had less antigen. Effectiveness results were largely unchanged when removing these data. In contrast to the primary analyses, efficacy estimates including only modern OCVs did not demonstrate any waning over the first five years (Figure S7). Finally, with the exception of one study,^29^ all effectiveness studies used a case-control design, most with notable risk of bias in terms of the ascertainment of exposure (usually through self-report) and in the comparability between cases and controls. Future evidence using more rigorous prospective designs, including rigorous vaccination ascertainment, longer duration of follow-up post-vaccination, and causal inference methods, may help improve confidence in estimates and potentially allow for regulatory approvals without the need for randomized trials.^40^ Our review was not able to address several pertinent policy-relevant questions related to vaccine use, including the optimal timing between doses, and when, and if, vaccination with one dose should be followed up with the full two-dose schedule. These decisions should consider not only the direct protection from the vaccine, as described in our analyses, but other epidemiological and operations considerations to ensure doses will have an impact.

Our pooled estimates of effectiveness were higher than those of efficacy over the first two years after vaccination. While the reasons for this are likely multifactorial, we hypothesize that this is in part explained by the fact that the RCTs were largely conducted in highly endemic communities, where the average age of infection is lower than in areas that are less exposed to cholera. In contrast, much of the observational data has come from less endemic communities. The median percentage of cases under five years of age in randomized trials was 27.8% compared to 16.7% in observational studies (Figure S8). Given that kOCV provides less protection for young children when compared to older individuals, we could expect to have lower estimates of protection in zones with a higher proportion of young children as cases. Another potential source for this discrepancy, which should be further explored, is the potential impact of indirect effects in the estimates of effectiveness.^41^

More than 31 countries have used kOCVs as part of their efforts to control cholera in the past decade,^42^ yet the lack of clear summaries on the duration of protection from one and two doses, combined with the global kOCV shortage, have complicated efforts to sustain vaccine-derived protection in communities at the highest risk. Our results provide a synthesis of the evidence on protection from kOCV studies since the mid-1980s, reconfirming that 2 doses of the vaccine provide moderate-to-high levels of protection at least into the 5th year after vaccination and that one dose can provide robust protection over at least the first two years after vaccination. This evidence supports the current policy of using one dose in outbreak response, where short-term protection is most critical. It is unclear if this level of protection would be sustained in populations that are largely immunologically naïve and whether a single dose strategy is efficient, particularly in endemic areas with high ongoing risk for cholera.^43^ Our review illustrates the temporal scale of waning direct protection, but indirect protection may wane even quicker in some settings due to human mobility patterns and the initial coverage of kOCV campaigns.^3,44^ While the world waits for universal access to water and sanitation in addition to better and more cholera vaccines, we must focus on improving the quality of vaccination campaigns in addition to improving surveillance of clinical cases and vulnerability factors so that we can dynamically track and respond to cholera risk.

## Disclaimer

The findings and conclusions in this report are those of the authors and do not necessarily represent the views of the Centers for Disease Control and Prevention.

## Data Availability

All data produced are available online at https://github.com/HopkinsIDD/kOCV-review.

