## Supplementary Material for "Protection from Killed Whole-Cell Cholera Vaccines: A Systematic Review and Meta-Analysis"

### *Supplementary Appendix*

**Table S1 Search terms and search results.** We did two rounds of search in five databases. The first round of search was on January 22, 2023. We did a second round of search on March 8, 2024 to include studies published between the two searches.

| **Engine** | **Language restriction** | **Exact Search Query** | **Number of records on Jan 22, 2023** | **Number of records on Mar 8, 2024** |
|---|---|---|---|---|
| Pubmed | None | cholera*[Title/Abstract] AND (vaccin*[Title/Abstract]) AND (effect*[Title/Abstract] OR efficacy[Title/Abstract] OR protect*[Title/Abstract] ) | 577 | 98 |
| Embase | None | cholera*:ab,ti AND vaccin*:ab,ti AND (efficacy:ab,ti OR effect*:ab,ti OR protect*:ab,ti) | 637 | 144 |
| Scopus | None | TITLE-ABS(cholera*) AND TITLE-ABS(vaccin*) AND TITLE-ABS(efficacy OR effect* OR protect*) AND NOT INDEX (embase) | 146 | 119 |
| ISI Web of Science | None | TI=(cholera* AND vaccin*) AND TS=(efficacy OR effect* OR protect*) | 227 | 24 |
| Cochrane Review Library | None | cholera* AND vaccin* AND (efficacy OR effect* OR protect*) | 0 | 9 |

**Table S2 Risk of bias summary for included observational studies. The maximum number of stars (indicating lowest risk of bias) is indicated next to MAX for each criterion. Assessment is based on the Newcastle-Ottawa Scale for observational studies** [1]**.**

| **CASE-CONTROL STUDIES** | **SELECTION MAX ****** | **COMPARABILITY MAX **** | **EXPOSURE MAX ***** |
|---|---|---|---|
| Wierzba et al. 2015 | *** | * | ** |
| Ivers et al. 2015 | **** | * | ** |
| Luquero et al. 2014 | **** | ** | * |
| Franke et al. 2018 | **** | * | * |
| Ferreras et al. 2018 | **** | * | * |
| Grandesso et al. 2019 | *** | | * |
| Sialubanje et al. 2022 | **** | * | |

| | | | |
|---|---|---|---|
| Malembaka et al. 2023 | **** | | * |
| Matias et al. 2023 | **** | * | * |
| **CASE-COHORT STUDIES** | **SELECTION MAX ****** | **COMPARABILITY MAX **** | **OUTCOME MAX ***** |
| Azman et al. 2016 | **** | * | ** |

**Table S3. Point estimates of two-dose efficacy or effectiveness of whole-cell OCV included in the meta-analysis.**

| Estimate type | Follow-up duration group | Location | Actual follow-up time (months) | VE (95% CI) | Study |
|---|---|---|---|---|---|
| Efficacy | 0-12 months | Bangladesh | [0, 12] | 47 (17-66) | **Ali et al, 2021** [2] |
| | | India | [0, 12] | 40 (-10-67) | Bhattacharya et al, 2013 [3] |
| | | Vietnam | [8, 10] | 66 (46-79) | Trach et al, 1997 [4] |
| | | Bangladesh | [1, 12] | 53 (38-66) | van Loon et al, 1996 [5] |
| | | Bangladesh | [0, 12] | 49 (10-71) | Qadri et al, 2015 [6] |
| | 12-24 months | Bangladesh | [12, 24] | 68 (42-82) | **Ali et al, 2021** [2] |
| | | India | [12, 24] | 72 (42-87) | Bhattacharya et al, 2013 [3] |
| | | Bangladesh | [12, 24] | 57 (42-70) | van Loon et al, 1996 [5] |
| | | Bangladesh | [12, 24] | 60 (23-79) | Qadri et al, 2015 [6] |
| | 24-36 months | Bangladesh | [24, 36] | 25 (-13-51) | **Ali et al, 2021** [2] |
| | | India | [24, 36] | 57 (26-75) | Bhattacharya et al, 2013 [3] |
| | | Bangladesh | [24, 36] | 42 (18-62) | van Loon et al, 1996 [5] |
| | 36-48 months | Bangladesh | [36, 48] | 48 (16-67) | **Ali et al, 2021** [2] |
| | | India | [36, 48] | 60 (33-76) | Bhattacharya et al, 2013 [3] |
| | | Bangladesh | [36, 48] | -28 (-114-31) | van Loon et al, 1996 [5] |
| | 48-60 months | India | [48, 60] | 81 (42-94) | Bhattacharya et al, 2013 [3] |
| Effectiveness | 0-12 months | Zambia | [0, 6] | 81 (72-84) | **Sialubanje et al, 2022** [7] |
| | | Malawi | [0, 3] | 83 (21-96) | **Grandesso et al, 2019** [8] |
| | | Haiti | [2, 12] | 84 (53-95) | **Franke et al, 2018** [9] |
| | | Haiti | [6, 14] | 87 (32-98) | Ivers et al, 2015 [10] |
| | | Guinea | [0, 5] | 87 (57-96) | Luquero et al, 2014 [11] |

| | | | | | |
|---|---|---|---|---|---|
| | 12-24 months | Haiti | [12, 24] | 66 (34-82) | **Franke et al, 2018** [9] |
| | | Haiti | [14, 22] | 64 (10-86) | Ivers et al, 2015 [10] |
| | | Haiti | [10, 27] | 69 (-71-94) | Matias et al, 2023 [12] |
| | | DRC | [12, 17] | 58 (27-76) | **Malembaka et al. 2024** [13] |
| | 24-36 months | Haiti | [24, 36] | 73 (30-90) | **Franke et al, 2018** [9] |
| | | India | [23, 34] | 69 (14-89) | Wierzba et al, 2015 [14] |
| | | DRC | [24, 36] | 25 (-19, 52) | **Malembaka et al. 2024** [13] |
| | 36-48 months | Haiti | [36, 48] | 94 (56-99) | **Franke et al, 2018** [9] |

**Table S4. Point estimates of one-dose efficacy or effectiveness of whole-cell OCV included in the meta-analysis.** The two estimates of one-dose OCV efficacy (Qadri et al. 2016 [15] and Qadri et al. 2018 [16]) were not included in the meta-analysis.

| Estimate Type | Follow-up duration group | Location | Actual follow-up duration (months) | VE (95%CI) | Study |
|---|---|---|---|---|---|
| Efficacy | 0-6 months | Bangladesh | [0, 6] | 58 (24-76) | Qadri et al, 2018[16] |
| | 6-12 months | Bangladesh | [6, 12] | 37 (-20-67) | Qadri et al, 2018[16] |
| | 12-18 months | Bangladesh | [12, 18] | 62 (34-78) | **Qadri et al, 2018** [16] |
| | 18-24 months | Bangladesh | [18, 24] | 67 (30-84) | Qadri et al, 2018[16] |
| Effectiveness | 0-6 months | Zambia | [0, 2] | 89 (43-98) | **Ferreras et al, 2018** [17] |
| | | Malawi | [0, 3] | 89 (36-98) | **Grandesso et al, 2019** [8] |
| | | Guinea | [0, 5] | 43 (-84-82) | Luquero et al, 2014 [11] |
| | | South Sudan | [0, 2] | 87 (70-100) | Azman et al, 2016 [18] |
| | 6-12 months | Haiti | [2, 12] | 92 (66-98) | **Franke et al, 2018** [9] |
| | 12-18 months | Haiti | [12, 24] | 40 (-31-73) | **Franke et al, 2018** [9] |
| | | DRC | [12, 17] | 53 (31-67) | **Malembaka et al. 2024** [13] |

| | | | | | |
|---|---|---|---|---|---|
| | | Haiti | [6, 22] | 67 (-62-93) | Ivers et al, 2015 [10] |
| | 24-30 months | DRC | [24, 36] | 46 (26-60) | **Malembaka et al. 2024** [13] |
| | | India | [23, 34] | 32 (-318-89) | Wierzba et al, 2015 [14] |

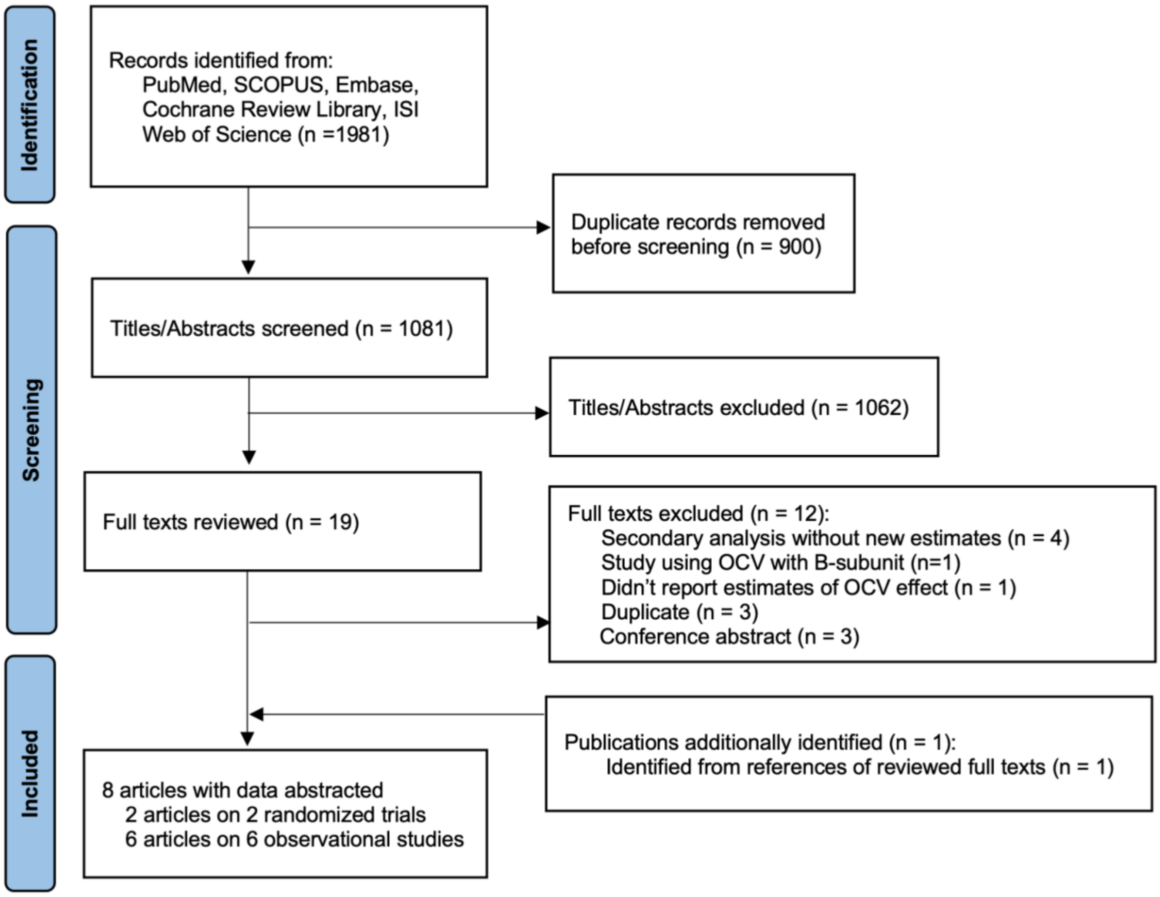


**Figure S1. PRISMA flow chart of screening process of newly identified records in 2024.** This flow chart illustrates the screening process for records identified in the new search, with date of publication restricted to January 1st, 2016 to March 8th, 2024.

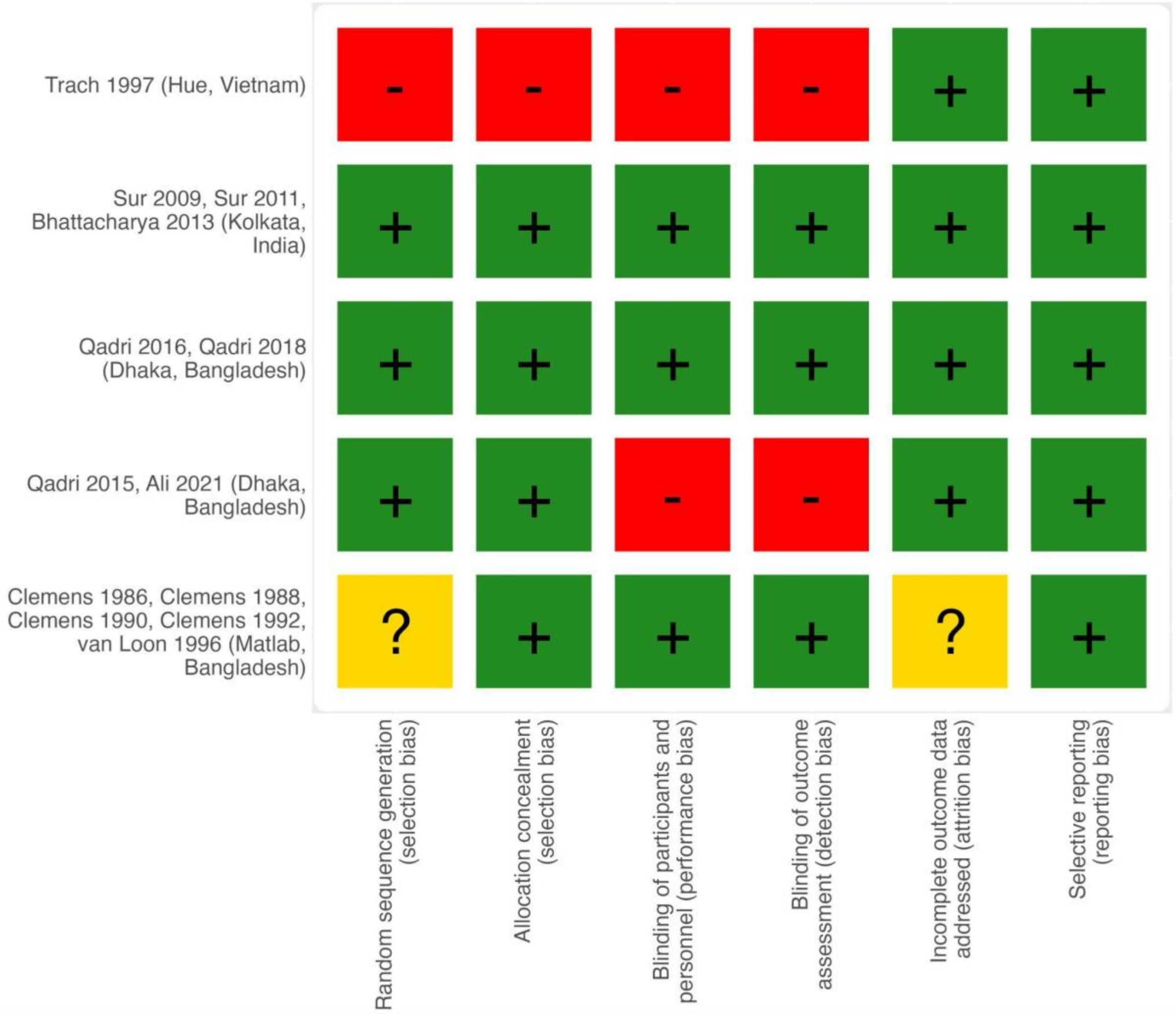


**Figure S2 Risk of bias summary for clinical trials following the Cochrane Collaboration Tool. Green cells represent low risk of bias, yellow cells indicate unclear risk of bias and red cells indicate high risk of bias.**

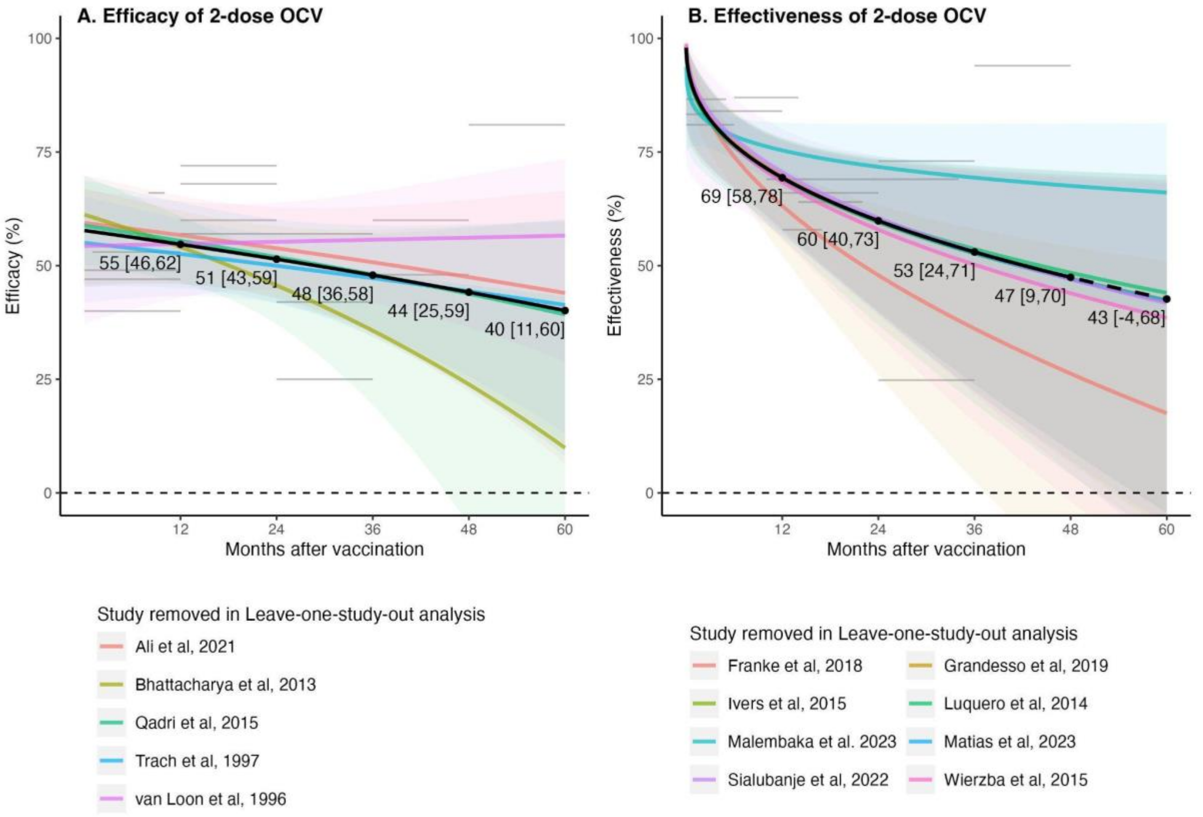


**Figure S3 Efficacy (A) and effectiveness (B) of two-dose kOCVs from leave-one-study-out analysis.** The black solid lines and labels are estimated efficacy or effectiveness using the full dataset of all two-dose estimates. The colored lines represent estimated efficacy and effectiveness after leaving out estimates from one study. The horizontal gray lines represent the full dataset that was used to fit the meta-regression models, the length of the line indicates the duration of follow-up (months since vaccination). The line's position on the y-axis marks the magnitude of the point estimate (%). The dashed horizontal line at y=0 denotes no protective effect (0%) of kOCV.

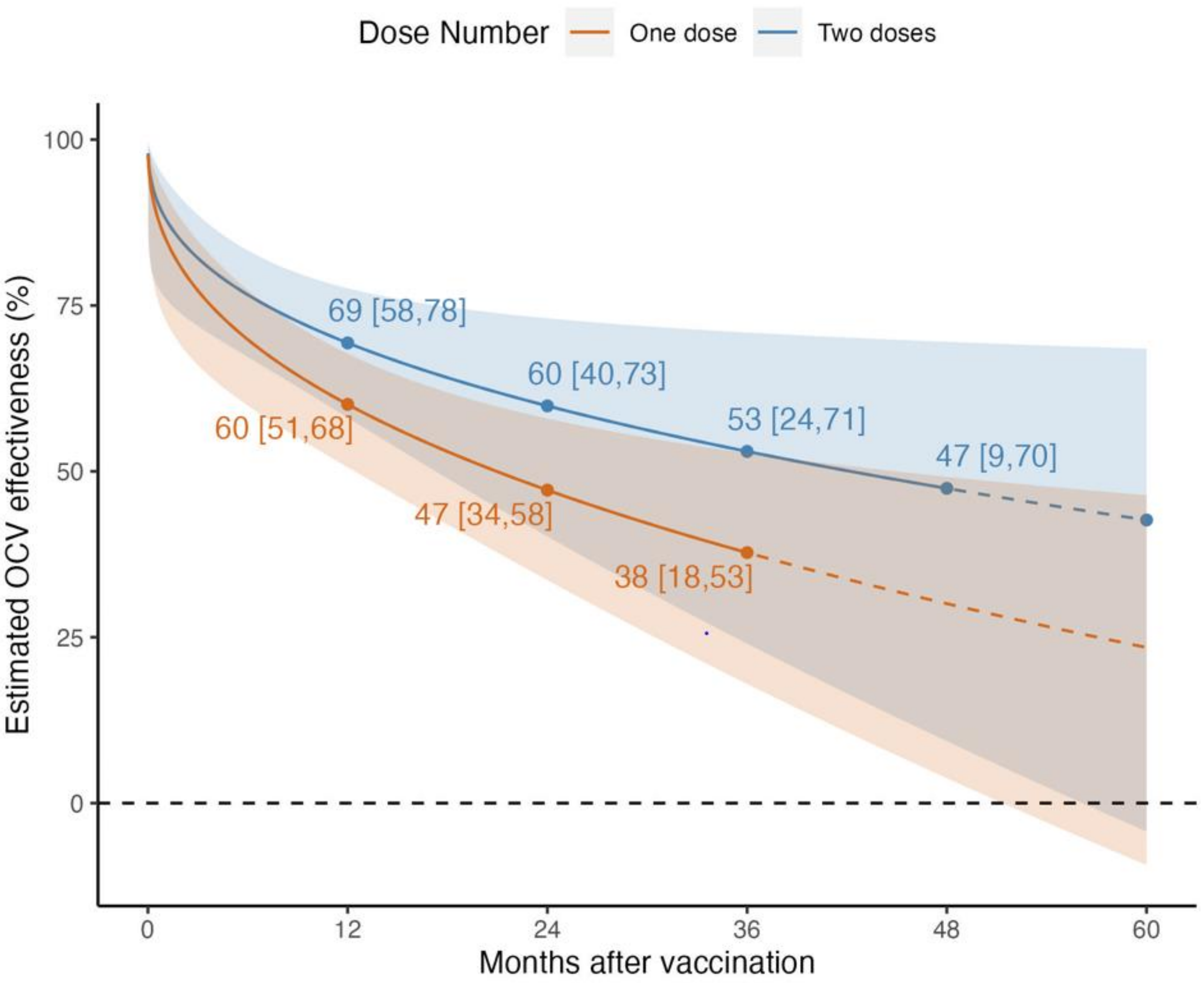


**Figure S4. Comparison of estimated effectiveness of one-dose and two-dose kOCV over time since vaccination predicted by meta-regression models.** The orange and pale blue curves represent effectiveness over time of one-dose and two-dose kOCV predicted by the meta-regression models, with the shaded bands representing the 95% confidence intervals and 95% prediction intervals. The filled circles indicate the predicted estimates at 12, 24, 36, 48 (for two-dose estimate only) months post-vaccination, the value and 95% confidence interval is labelled below. The dashed horizontal line denotes no protective effect (0%) of kOCV. The dashed curves represent the extrapolated effectiveness for the follow-up period without any reported data from literature.

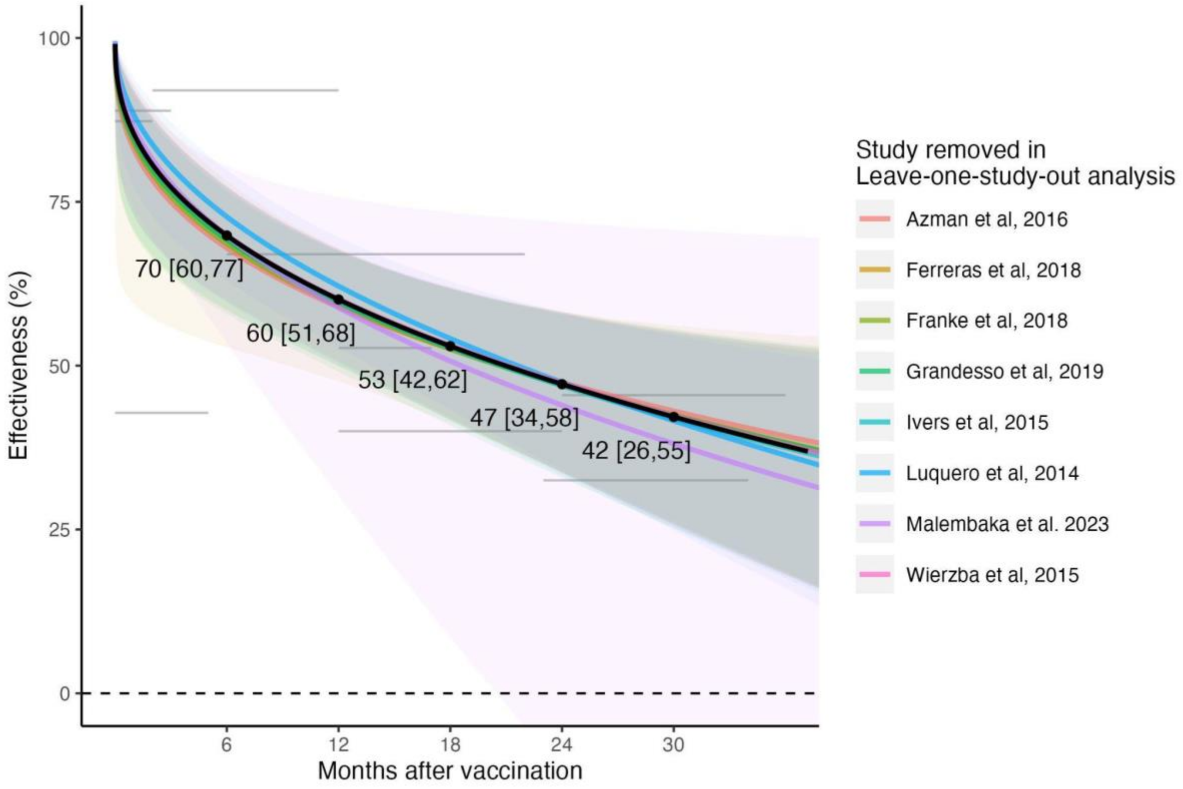


**Figure S5 Effectiveness of one-dose kOCVs from leave-one-study-out analysis.** The black solid lines and labels are estimated efficacy or effectiveness using the full dataset of all two-dose estimates from included studies. The colored lines represent estimated efficacy and effectiveness after leaving out estimates from one study. The horizontal gray lines represent the full dataset that was used to fit the meta-regression models, the length of the line indicates the duration of follow-up (months since vaccination). The line's position on the y-axis marks the magnitude of the point estimate (%). The dashed horizontal line at y=0 denotes no protective effect (0%) of kOCV.

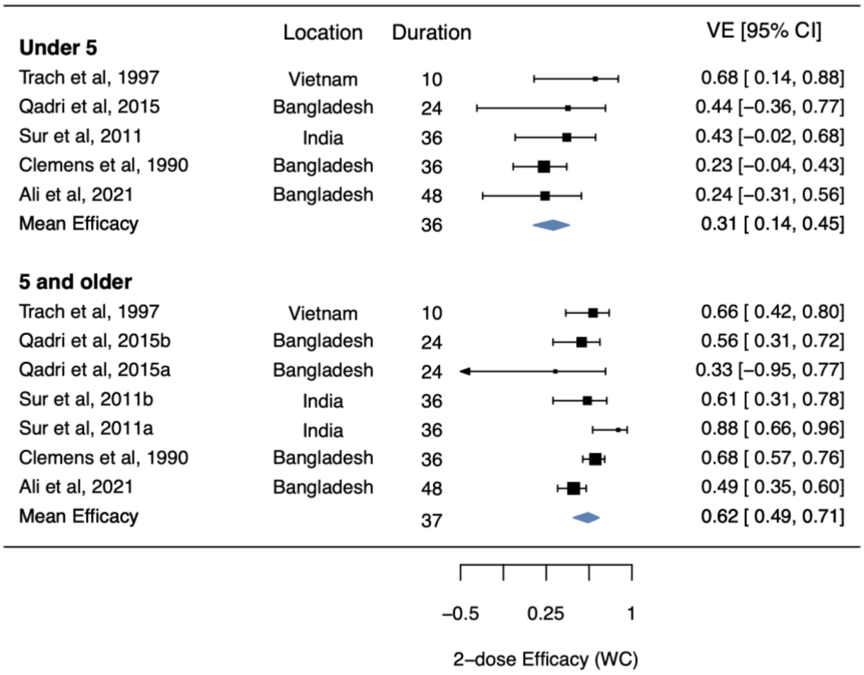


**Figure S6. Pooled efficacy of two-dose whole-cell kOCV by age group.** Duration of the mean efficacy of each age group was weighted mean duration of the included estimates. Clemens et al. 1990 [19] was using three doses instead of two doses but is still included in the analyses. Sur et al. 2011a and Qadri et al. 2015a were subgroup estimates for participants aged between 5 and 15, while Sur et al. 2011b and Qadri et al. 2015b were subgroup estimates for participants aged above 15 years old [6,20]. The estimates included in this pooled analyses were efficacy estimates for the whole follow-up period. Black bars and squares show 95% confidence intervals and point estimates of efficacy for the studies. Blue diamonds show the pooled efficacy estimates for participants under 5 or 5 and older.

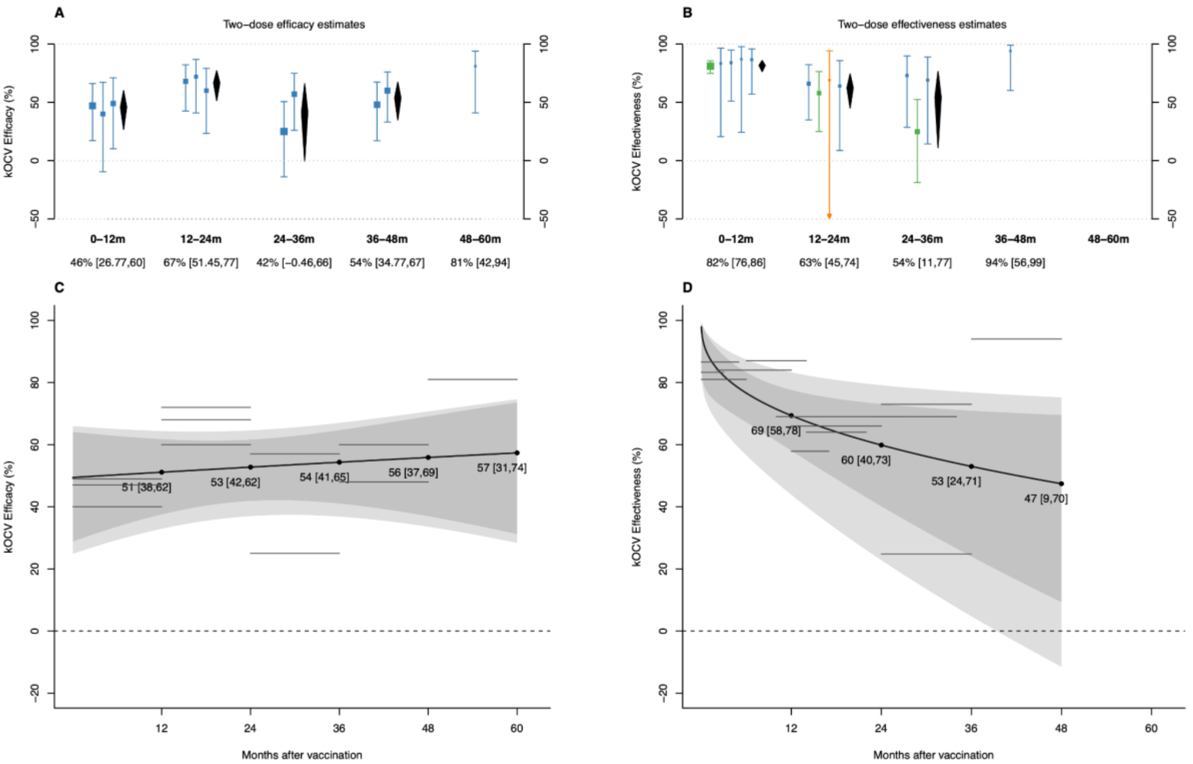


**Figure S7. Stratified and meta-regression estimates of the efficacy and effectiveness of two doses of killed whole-cell OCV (kOCV) as a function of time since vaccination, after removing vaccines that came before Shanchol vaccines.** The upper panels illustrate stratified estimates of efficacy (A) and effectiveness (B) by time since vaccination bin (0-12, 12-24, 24-36, 36-48, 48-60 months after vaccination). The trial conducted in Matlab, Bangladesh [5] and the trial conducted in Hue, Vietnam [4] were not included in this figure and meta-regression analysis as they used pre-Shanchol vaccines. Estimates are grouped into the five follow-up duration categories by the midpoint of the time window during which the estimate was measured. Bars and squares show 95% confidence intervals (CI) and point estimates of efficacy or effectiveness for each literature, colored by vaccine type (blue: “Shanchol; green: Euvichol-plus; orange: Euvichol). Diamonds in black show the estimated average efficacy or effectiveness and 95% CI by follow-up period, with numerical values shown at the bottom of the x-axis in black. If there is only one estimate in the follow-up period, the estimate from the study is presented on the x-axis. The bottom panels illustrate meta-regression results for average two-dose (A) efficacy and (B) effectiveness as a function of time since vaccination, with the shaded envelope representing the 95% confidence intervals and 95% prediction intervals. The horizontal gray lines represent the data from the literature that were used to fit the meta-regression models, the length of the line indicates the duration of follow-up (months since vaccination). The line’s position on the y-axis marks the magnitude of the point estimate (%). The dashed horizontal line at y=0 denotes no protective effect (0%) of kOCV.

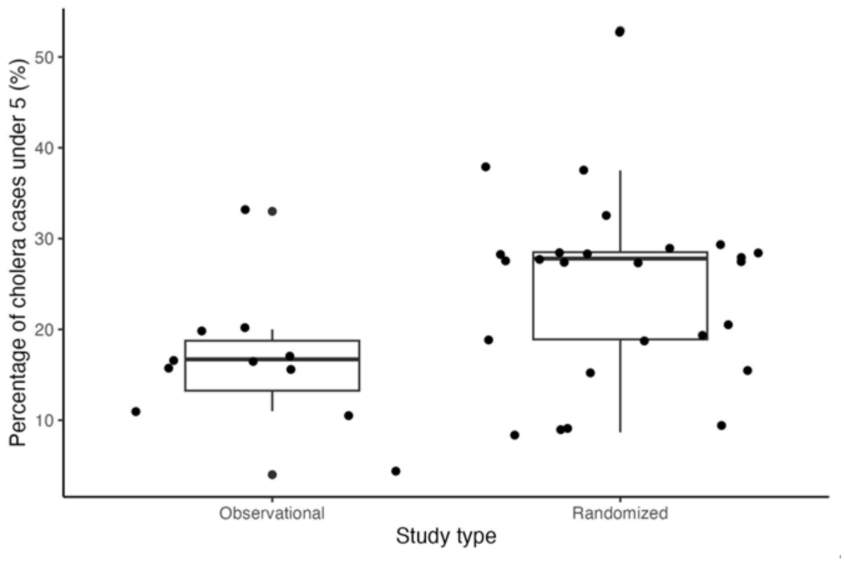


**Figure S8. Distribution of percentage of cholera cases under 5 years old in efficacy studies and effectiveness studies.** Each point represents the percentage of cholera cases that are under 5 years old in one estimate. The box demarcates the 25th, 50th (median) and 75th percentile and the whiskers extend beyond the 15th and 75th percentiles by 1.5 times the interquartile range. The median percentage of cases under 5 is 16.7% for observational (effectiveness) studies and 27.8% for randomized (efficacy) studies.

13. Malembaka EB, Bugeme PM, Hutchins C, Xu H, Hulse JD, Demby MN, et al. Effectiveness of one dose of killed oral cholera vaccine in an endemic community in the Democratic Republic of the Congo: a

matched case-control study. Lancet Infect Dis. 2024. doi:10.1016/S1473-3099(23)00742-9
